# Prevalence of Salivary IgA Reacting with SARS-CoV-2 among Japanese People Unexposed to the Virus

**DOI:** 10.1101/2022.01.09.22268986

**Authors:** Keiichi Tsukinoki, Tetsuro Yamamoto, Jiro Saito, Wakako Sakaguchi, Keiichiro Iguchi, Yoshinori Inoue, Shigeru Ishii, Chikatoshi Sato, Mina Yokoyama, Yuki Shiraishi, Noriaki Kato, Hiroyasu Shimada, Akio Makabe, Akihiro Saito, Masanori Tanji, Isao Nagaoka, Juri Saruta, Tetsutaro Yamaguchi, Shigenari Kimoto, Hideyo Yamaguchi

## Abstract

While the COVID-19 pandemic caused by SARS-CoV-2 has posed a threat to public health as the number of cases and COVID-19-related deaths are increasing worldwide, the incidence of the virus infection are extremely low in Japan compared with many other countries. To explore the reason for this strange phenomenon, we hypothesized the high prevalence of “natural” secretory IgA in saliva as mucosal IgA reacting with SARS-CoV-2, and thus surveyed the positivity for, as well as levels of, such reactive salivary IgA in a cohort of Japanese people of a wide range of age. The major findings were that 95/180 (52.78 %) of overall individuals who had not been exposed to SARS-CoV-2 were positive for salivary IgA with the levels ranging from 0.002 to 3.272 ng/ml, and that there may be a negative trend in positivity for salivary IgA according to age. These results suggest a role of mucosal IgA in host defense against SARS-CoV-2 infection.

**One Sentence Summary:** “Natural” secretory immunoglobulin A autoantibodies may play a role in mucosal defense against SARS-CoV-2.

## INTRODUCTION

In December 2019, a novel coronavirus termed SARS-CoV-2 was identified as the cause of a new pandemic of the severe acute respiratory syndrome known as COVID-19 (1). Since then, this emerging virus infection has globally spread with an unexpectedly high speed and has created a public health crisis, as well as the major economic and social burdens. As of November 18 2020, the ongoing pandemic of COVID-19 has infected more than 56 million people worldwide with a mean population-based infection rate (percentage of numbers of PCR-confirmed cases per whole population) being 0.721 % (2). This is thought to be due to the highly efficient person-to-person transmission of SARS-CoV-2 and lack of population-level immunity (3). However, incidence of COVID-19 markedly varied by geographical areas and nations. For example, infection rates as of November 1 2020 for G7 Countries comprising USA, France, UK, Italy, Germany, Canada and Japan were estimated to be 2.802 %, 2.176 %, 1.524 %, 1.173 %, 0.651 %, 0.638 %, and 0.081 %, respectively (4). The data indicate that Japan remained one of the countries least severely affected by the ongoing COVID-19 pandemic owing to such an extremely low incidence of the disease, despite that none of Japanese people had not yet undergone any vaccination for COVID-19. At this moment, there is no rational explanation for this curious phenomenon.

Several recent reports suggest that immunoglobulin G (IgG) responses and, to a greater extent, immunoglobulin A (IgA) responses to SARS-CoV-2 play a role in protecting the human host through neutralization of the virus infectivity (5-9). IgA is by far the most abundant class immunoglobulin (Ig) in humans, each day IgA is mostly produced by plasma cells locally existing in mucosa-associated lymphoid tissues in amounts of 3 - 5 g that exceeds the combined production of all other isotypes (10,11). Compared to other Ig classes, IgA has unique structural properties in terms of differences in its glycosylation patterns and molecular forms, as well as the presence of more than one receptor due to its dimeric or more highly polymeric forms when existing in mucosal secretions (12-14). In humans, the most common IgA form present in serum is a monomer, whereas at mucosal sites, it is produced as polymeric molecules, foremost as dimeric IgA that consists of two IgA molecules, linked by an N-glycosylated protein called joining-chain (J-chain) (12). The polymeric Ig receptor (pIgR), which is expressed on the basolateral pole of epithelial cells, binds to the J-chain and releases IgA into the lumen, such as the respiratory tract and the oral cavity, as secretory IgA (sIgA) (14,15). It is well accepted that sIgA constitutes a principal component of mucosal immunity and thus it is referred to as mucosal IgA.

Recently, Wang and his coworkers characterized the IgA response to SARS-CoV-2 in a cohort of convalescent individuals after diagnosis with COVID-19 (8). The results showed that plasma (serum) IgA antibodies specific to SARS-CoV-2 antigens are two-fold less potent than IgG equivalents, but that the virus-specific sIgA antibodies existing in the mucosal secretions mostly as dimers are more than one log more potent than their monomeric counterparts present in the serum against SARS-CoV-2. Thus, the authors concluded that sIgA responses may be particularly valuable for protection against SARS-CoV-2 infection and for vaccine efficacy (8).

SARS-CoV-2 primarily infects the mucosal surfaces of the nasopharynx and the upper respiratory tract (16,17), as well as the oral cavity (18), at least until advanced stages of the disease (COVID-19) when viral RNA may become detectable in the circulation. The virus also infects the glands and mucosae of the oral cavity which harbor epithelial cells expressing angiotensin converting enzyme 2 (ACE2) and several other receptors for the virus spike (S) proteins, particularly the receptor binding domain (RBD) (18-20). These findings also underscore a crucial role of sIgA in protecting mucosal surfaces against SARS-CoV-2 by neutralizing the virus and/or impeding its attachment to epithelial cells at the initial stage of the virus infection.

There is recent evidence showing that anti-SARS-CoV-2 immunity not only occurs after a natural infection, but may also precede such an active infection. Cross-reactivity to SARS-CoV-2 antigen peptides has been identified on T-cells and B-cells from pre-pandemic donors; S protein-reactive CD4+ T-cells are not only detected in a large fraction of patients with COVID-19 but also in a smaller but considerable fraction of the healthy individuals with no history of COVID-19 (21,22). Consistent with this, it was also demonstrated that antibodies, probably including polyreactive natural autoantibodies (23), cross-reacting with SARS-CoV-2 have been detected in healthy individuals unexposed to the virus (9,24,25), and that a cross-reactive human IgA monoclonal antibody, which binds to SARS-CoV-2 S proteins with resulting in competitive inhibition of ACE2 receptor binding, has been successfully developed (26).

All these findings led us to the hypothesis that lower population-based susceptibility to SARS-CoV-2 infection seen in Japan compared to G7 and many other countries is associated with higher prevalence of sIgA reactive with the virus among Japanese people. Therefore, as an initial step to approach this possibility, we implemented a cohort study with healthy Japanese participants unexposed to SARS-CoV-2, in which we measured concentrations of the virus-reactive mucosal IgA (sIgA) using saliva samples because sIgA, as a secreted antibody, is noninvasively accessible in saliva (27,28). Measurements were also performed for SARS-CoV-2-reactive IgA and IgG concentrations in serum for comparison.

## RESULTS

Consent to the present study was provided and the questionnaire was completed by 139 adult participants (aged ≥20 years) themselves or by each legal representative of 41 juvenile participants (aged <20 years). Thus, both saliva and blood samples and only saliva samples were collected from all the enrolled adults and juveniles, respectively. All of the 180 participants had no clinical history nor current status consistent with COVID-19 and tested negative for SARS-CoV-2 by RT-PCR.

All of the 180 individuals were tested whether they had detectable amounts of SARS-CoV-2-reactive salivary IgA or not. The results showed that among them 95 individuals (52.78 %) tested positive as shown in Table1. The fraction that tested positive did not differ with gender, but varied by age; higher and lower positivities were observed for the youngest age group (<20 years) and the oldest age group (≥51 years), respectively, compared to the two intermediate age groups (20-35 years and 36-50 years groups). Thus, it was suggested that there is a negative trend of positivity for saliva IgA according to age. No association was found for salivary IgA between test results and comorbidities, history of any vaccinations or daily oral care and/or dietary habits (data not shown).

We observed a broad range of reactive salivary IgA concentrations with an over 1,600-fold difference between the extremes (0.002 and 3.272 ng/ml) among the 95 test-positive individual samples. However, the results of 90 of the 95 samples (94.7 %) clustered around a median of 0.102 ng/ml (Fig.1). Similar distribution patterns and medians of levels for test-positive samples were also observed in each age group; medians for <20 years-, 20-35 years-, 36-50 years- and >50 years-groups were 0.111, 0.134, 0.067 and 0.121 ng/ml, respectively. Three test-positive samples with exceptionally high reactive salivary IgA levels between 1.03 and 3.27 ng/ml were derived from individuals in the 20-35 years-group (1 sample) or the 26-50 years-group (two samples).

**Fig. 1.**
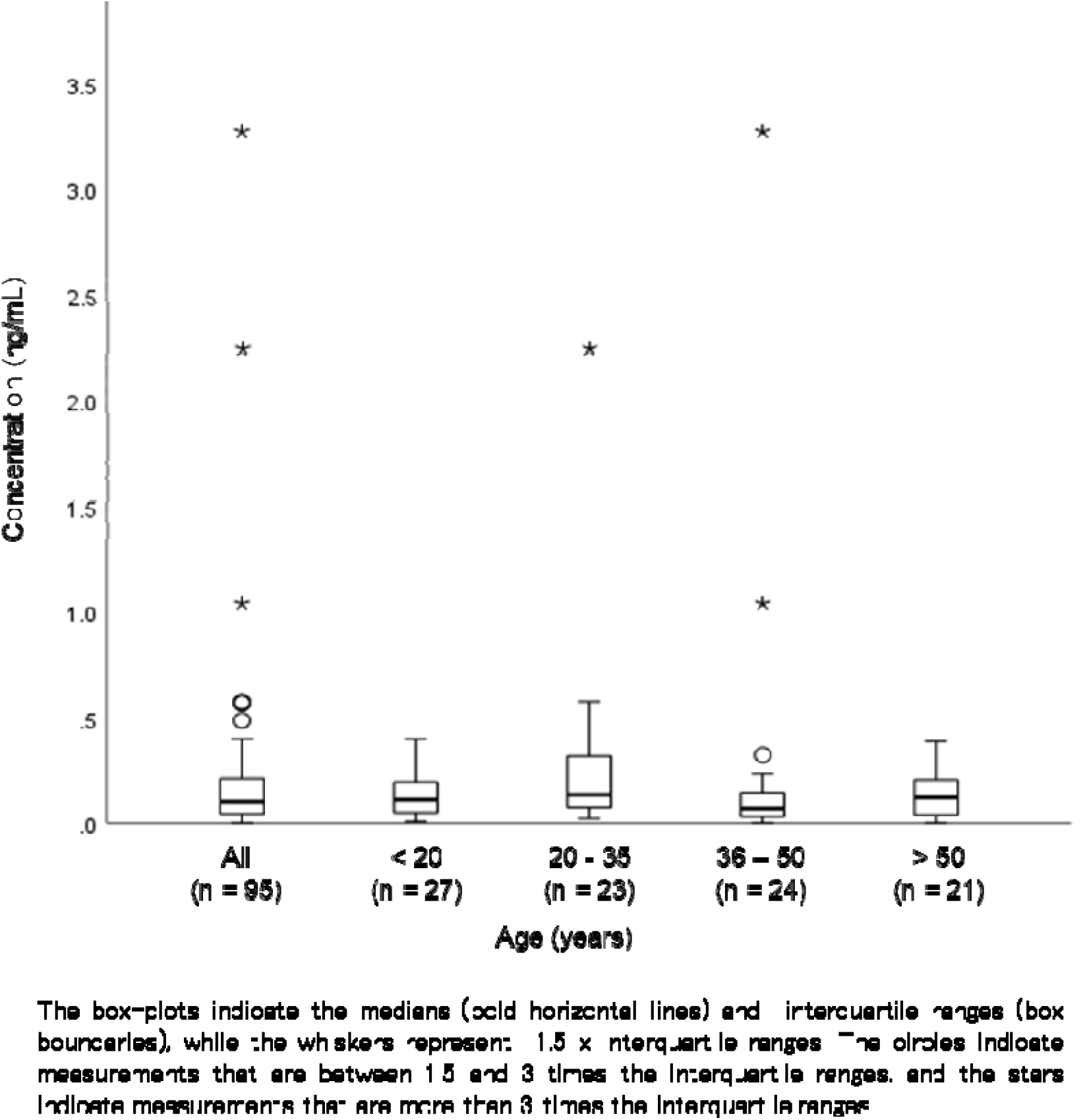
Presence and levels of salivary IgA reactive with SARS-CoV-2 in all participants and fractionated age groups of the study cohort.

One hundred and thirty-nine adult participants provided matched saliva and serum samples collected during the same visit. All samples were tested multiplex assay for measuring levels of SARS-CoV-2-reactive IgA in saliva, as well as those of equivalent IgA and IgG in serum. Sixty-eight (48.92 %), 24 (17.27 %) and 17 individuals (12.23 %) were positive for SARS-CoV-2-reactive salivary IgA, serum IgA and serum IgG, respectively (Table 2). The fraction that tested positive for each Ig equivalent varied by age and BMI. A negative trend of test positivity according to age and BMI were observed for serum IgG and salivary IgA, respectively, while the proportion of test positivity for serum IgA and serum IgG in the fraction of BMI at a reference interval (20-25) was higher than the other two BMI fractions (Table 2).

**Table 1.**
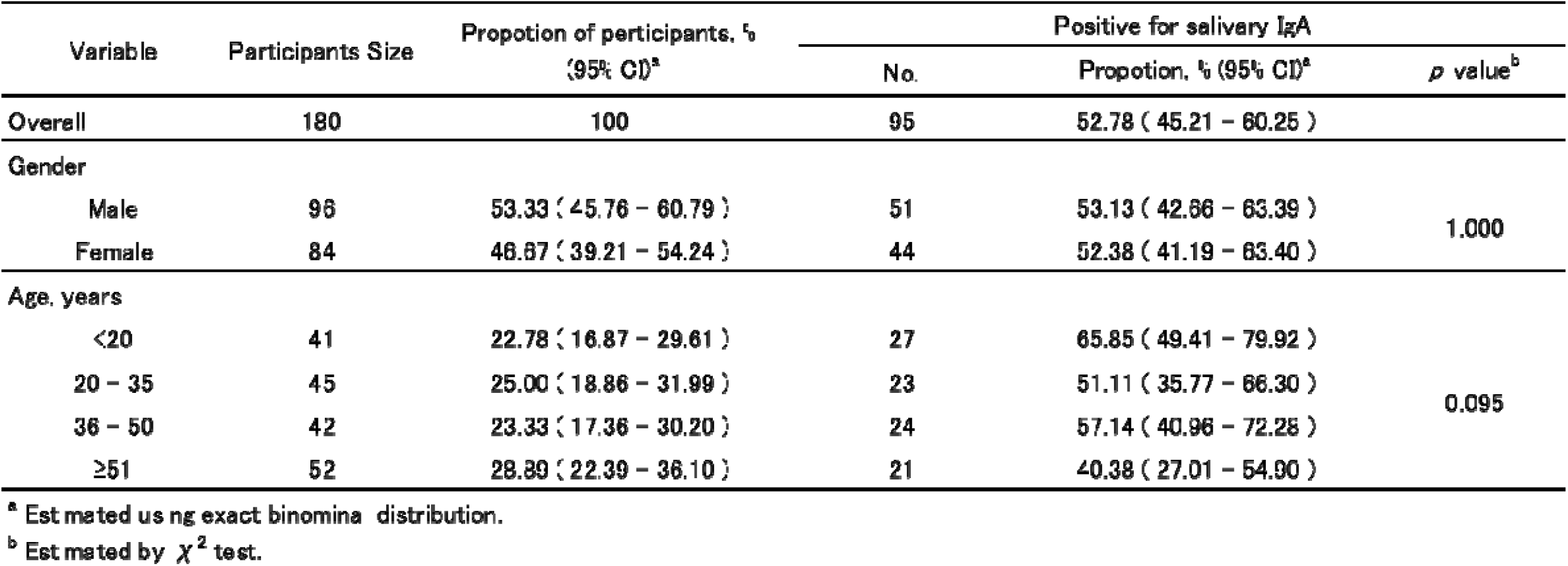
Proportion positive for salivary IgA reactive with SARS-CoV-2 in all participants of the study cohort stratified by gender and age groups.

**Table 2.** Proportion positive for salivary IgA, Serum IgA and serum IgG reactive with SARS-CoV-2 in an adult participant group (age ≥20 years) of the study cohort stratified by gender, age and BMI subgroups.

| Variable | Participant<br>s<br>size | Propotion of participants, %<br>(95% CI)<sup>a</sup> | Positive for salivary IgA |  |  | Positive for serum IgA |  |  | Positive for serum IgG |  |  |
| --- | --- | --- | --- | --- | --- | --- | --- | --- | --- | --- | --- |
|  |  |  | No. | Propotion, % (95% CI)<sup>a</sup> | *p* value<sup>b</sup> | No. | Propotion, % (95% CI)<sup>a</sup> | *p* value<sup>b</sup> | No. | Propotion, % (95% CI)<sup>a</sup> | *p* value<sup>b</sup> |
| Overall | 139 | 100 | 68 | 48.92 ( 40.35 – 57.53 ) |  | 24 | 17.27 ( 11.39 – 24.59) |  | 17 | 12.23 ( 7.29 – 18.86 ) |  |
| Gender |  |  |  |  |  |  |  |  |  |  |  |
| Male | 71 | 51.08 ( 42.47 – 59.65 ) | 36 | 50.70 ( 38.56 – 62.78 ) | 0.735 | 13 | 18.31 ( 10.13 – 29.27 ) | 0.824 | 8 | 11.27 ( 4.99 – 21.00 ) | 0.799 |
| Female | 68 | 48.92 ( 40.35 – 57.53 ) | 32 | 47.06 ( 34.83 – 59.55 ) |  | 11 | 16.18 ( 8.36 – 27.10 ) |  | 9 | 13.24 ( 6.23 – 23.64 ) |  |
| Age, years |  |  |  |  |  |  |  |  |  |  |  |
| 20 – 35 | 45 | 32.37 ( 24.69 – 40.83 ) | 23 | 51.11 ( 35.77 – 66.30 ) | 0.259 | 10 | 22.22 ( 11.20 – 37.09 ) | 0.564 | 9 | 20.00 ( 9.58 – 34.60 ) | 0.055 |
| 36 – 50 | 42 | 30.22 ( 22.72 – 38.57 ) | 24 | 57.14 ( 40.96 – 72.28 ) |  | 6 | 14.29 ( 5.43 – 28.54 ) |  | 6 | 14.29 ( 5.43 – 28.54 ) |  |
| ≥51 | 52 | 37.41 ( 29.36 – 46.01 ) | 21 | 40.38 ( 27.01 – 54.90 ) |  | 8 | 15.38 ( 6.88 – 28.08 ) |  | 2 | 3.85 ( 0.47 – 13.21 ) |  |
| BMI |  |  |  |  |  |  |  |  |  |  |  |
| <20 | 31 | 22.30 ( 15.68 – 30.14 ) | 17 | 54.84 ( 36.03 – 72.68 ) | 0.337 | 2 | 6.45 ( 0.79 – 21.42 ) | 0.041 | 1 | 3.23 ( 0.08 – 16.70 ) | 0.099 |
| 20 – 25 | 90 | 64.75 ( 56.20 – 72.66 ) | 45 | 50.00 ( 39.27 – 60.73 ) |  | 21 | 23.33 ( 15.06 – 33.43 ) |  | 15 | 16.67 ( 9.64 – 26.00 ) |  |
| ≥25 | 18 | 12.95 ( 7.86 – 19.69 ) | 6 | 33.33 ( 13.34 – 59.01 ) |  | 1 | 5.56 ( 0.14 – 27.29 ) |  | 1 | 5.56 ( 0.14 – 27.29 ) |  |

<sup>a</sup> Estimated using exact binominal distribution.
<sup>b</sup> Estimated by $\chi^2$ test.

We also conducted correlation analysis to learn possible correlation of the test positivity for saliva IgA with that for serum IgA and/or serum IgG. The results showed no significant correlation with the value for either serum Ig equivalents (Table 3).

**Table 3.**
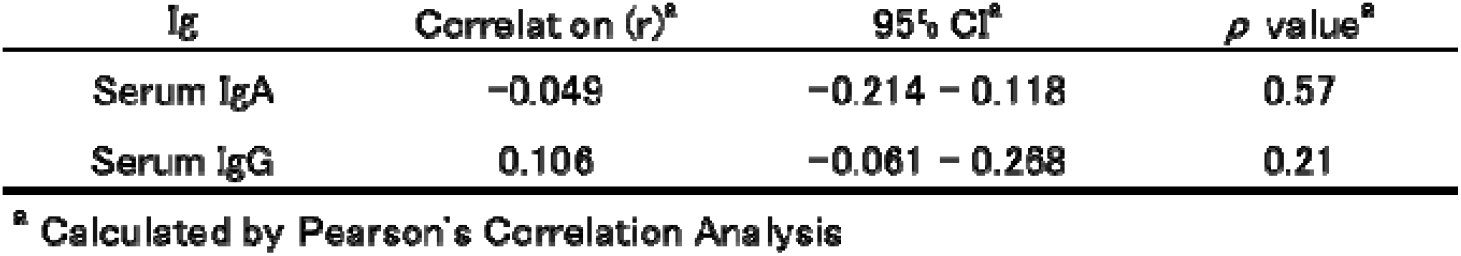
Correlation of the positivity for SARS-CoV-2 of salivary IgA with equivants of serum IgA and IgG in an adult participant group (aged 20 years; n = 139) of the study cohort.

## DISCUSSION

Our study aimed to survey the presence of sIgA in saliva, together with IgA and IgG in serum, capable of reacting with SARS-CoV-2 among Japanese people of a wide range of age from infants to the elderly who had been unexposed to the virus. The results evidenced the presence of salivary IgA reactive with a SARS-CoV-2 S1 antigen at detectable levels in a substantial fraction of enrolled individuals of all ages. Indeed, 95/180 (52.78 %) of overall individuals were positive for anti-SARS-CoV-2 salivary IgA. In addition, we found that there may be a negative trend in positivity for the virus-reactive saliva IgA according to age; the highest and the lowest prevalence of positivity were observed for the youngest age group (<20 years, 65.85 %) and the oldest age group (≥51 years, 40.38 %), respectively, compared to two intermediate aged adult groups.

Recently, a huge body of evidence has accumulated that susceptibility to SARS-CoV-2 infection generally increases with age. Compared to younger / middle-aged adults, children are less susceptible to the virus infection, while estimated susceptibility in older adults is considerably higher (29-37). Particularly, low fragility of children and adolescents against SARS-CoV-2 has become a matter of epidemiological and virological concern and intensive studies have been performed to identify the immune mechanisms implicated. As a result, several protective mechanisms including differences in the timing and nature of the induced cytokine responses and high amounts of ACE2 expressed, as well as trained immunity acquired from frequent viral infections and/or routine vaccinations, have been hypothesized (38). High positivity for SARS-CoV-2-reactive salivary IgA in children and adolescents, compared to adults, as observed in the present study may contribute to lower their susceptibility to the virus. So far as concerned with adult individuals, our results are in accordance with the data precedently presented by Tsukinoki and colleagues, who described a rate of positive SARS-CoV-2 cross-reactive saliva IgA as high as 46.7 % among the virus-uninfected professional workers of Kanagawa Dental University Hospital with being significantly lower in prevalence of positivity for individuals aged ≥50 years compared with those aged ≤49 years (39). The results of these two studies indicating that there is a decrease in SARS-CoV-2-reactive saliva IgA with increasing age are in line with preceding researches (40-42).

There are numerous reported human studies which indicated the association of low levels of or decreases in salivary IgA with incidence of upper respiratory infections, mostly common cold, in various study cohorts, such as infants (43), healthy children (44-47), children with adenoid hyperplasia (48), children with Down’s syndrome (49), elite athletes (50-53), individuals during intense physical training (54,55) and those who had undergone tonsillectomy or adenoidectomy in childhood (56). These results suggest that individuals who are deficient in salivary IgA, particularly those who are negative for salivary IgA reactive with SARS-CoV-2, may be at higher risk for the virus infection. In this context, unexpectedly high rates of SARS-CoV-2-reactive salivary IgA positivity seen in unexposed Japanese participants could be a part at least of the reason for extremely low incidence of COVID-19 in this country.

We also found that SARS-CoV-2-reactive antibodies are contained not only in salivary IgA but also in serum IgA and serum IgG. However, there was no correlation in the positivity or level of the virus-reactive antibodies between saliva IgA and both of the two Ig isotypes existing in serum. This may be probably due to different mechanisms involved in regulation of generation and/or dynamics of each type of Igs. It is well accepted that sIgA, a major component of salivary IgA, is generated and released at mucosal inductive site tissues, while the serum counterpart is, like IgG, derived from a distinct source, the bone marrow (57,58). Our postulate is that all of salivary IgA, serum IgA, and serum IgG antibodies reacting with SARS-CoV-2 might work co-operatively through binding to a common site RBD on S1 proteins of the virus, even though their binding affinities are different.

The present study has both strengths and limitations. By implementing this immunological survey in a community-based cohort study with a wide age range of healthy Japanese people who had been unexposed to SARS-CoV-2, we were able to collect preliminary but unprecedented epidemiological data regarding the prevalence and levels of assumable polyreactive natural salivary IgA autoantibodies that exhibit reactivity with the virus. To the best of our knowledge, our survey, along with the preceding one (38), is the only available study which has provided data useful for considering a putative role mucosal natural IgA antibodies in protecting the human host from SARS-CoV-2 infection. Supportably, the presence of natural polyreactive sIgA autoantibodies acting as the frontline of mucosal defense against various infections has been demonstrated (59).

Our study has several limitations. Firstly, our sample size was not large with limiting the robustness of our findings in saliva, as well as in serum. If such samples could be increasingly available, statical power for the analyses presented here will be increased. Secondly, we do not know whether and what levels of the salivary IgA detected in the presented study are protective through neutralization against SARS-CoV-2 infection. Thirdly, because of the lack of follow-up data, it was also unable to directly correlate negativity for or low levels of SARS-CoV-2-reactive salivary IgA with the feasibility to have the virus infection.

Nevertheless, the results obtained from the present study still lead us to favor the hypothesis that “natural” salivary IgA could contribute to lower the susceptibility to SARS-CoV-2 infection. According to the latestly publicized data, there is no significant difference in the completion rate of vaccination among the G7 Countries ranging from 60.1 % for USA to 77.7 % for Japan (as of December 6, 2021) (60). Despite that, the mean number of new COVID-19 cases per 1,000,000 people during the first week of December, 2021 for Japan was extremely low of 0.81 compared with the equivalents for other six G7 Countries as follows: 358.56 (USA); 686.26 (UK); 656.85 (France); 622.66 (Germany); 242.71 (Italy); and 83.79 (Canada) (4). Considering these global epidemiological data and our findings that a larger fraction of Japanese people are conferred with SARS-CoV-2-reactive salivary IgA, it is probable that mucosal immunity due to natural sIgA autoantibodies play a positive role in achieving herd immunity.

In conclusion, our preliminary results, obtained with SARS-CoV-2 unexposed people reflecting the general population in Japan, showed the presence of natural sIgA autoantibodies reactive with the virus at a considerably high rate. This may be related with low incidence of COVID-19 in Japan.

## MATERIALS AND METHODS

### Study participants

This epidemiological humoral immunity survey was a cross-sectional cohort study undergoing in the Kantoh District of Japan that is located in the middle area of the Main Island of the country constituted of Metropolitan Tokyo and three neighboring Prefectures (Kanagawa, Chiba and Saitama). This survey was conducted in two groups of individuals invited to participate in the present study. Group I volunteers aged between 20 and 75 years were recruited from the general public in the Kantoh District. Group II consisted of children and adolescents under treatment for their dental conditions, university students and dentists aged between 3 and 71 years in Kanagawa Dental University Hospital, Yokosuka, Kanagawa, all of whom were inhabitants of the Kantoh District. The inclusion criteria were: to have Japanese racial background; to have had no history or experience of diagnosed COVID-19, SARS-CoV-2 PCR-positive, nor COVID-19-related symptoms; and to have had not experienced common cold-like symptoms during the preceding two weeks. Individuals were excluded if: they have had received anti-SARS-CoV-2 vaccination; they were under treatment for systemic diseases or injuries; they had oral mucosal diseases with local bleeding; or they had participated in another clinical study within one month prior to the current study period.

### Study approval and ethics

Approval to undertake the study was obtained from the Kanagawa Dental University Research Ethics Review Board (approval number: 792) on April 6, 2021. The study adhered to all the guidelines set forth in the Declaration of Helsinki (amended in October 2013), the Ethical Guidelines for Medical Research in Humans (noticed by the Ministry of Health, Labor and Welfare, the Japanese Government, in December 2014 [amended in February 2017]) and the Guidance of the Ethical Guidelines for Medical Research in Humans (enacted by the Ministry of Education and Science and the Ministry of Health, Labor and Welfare, the Japanese Government, in February 2015 [amended in March 2017]). Written informed consent was obtained from all participants themselves or informed assent from their legal representatives, mostly parents (in the case of children or adolescent participants aged below 20 years), prior to the study onset.

The study was registered on April 1, 2021 in the database of the University Hospital Medical Information Network Clinical Trial Registry (UMIN-CTR) that meets the JCMJE standards. The study ID was UMIN000043717.

### Study design and procedures

All eligible participants were requested to visit one of the following four study centers: Medical Station Clinic (Tokyo); Kanagawa Dental University Hospital; Kanagawa Dental University Yokohama Clinic (Kanagawa); and Sato Dental Clinic (Kanagawa), on April 26 – August 1, 2021 for collection of both saliva and blood samples (in adult participants aged ≥20 years) or for collection of saliva samples only (in children or adolescent participants aged <20 years). All the participants were also instructed to visit the indicated study center early in the morning on the appointed day and after arrival, to refrain from taking any foods and drinks, as well as from tooth-brushing, for at least one hour before the collection of saliva samples, and then to take a cup of water for cleaning the oral cavity.

One hour later, saliva samples were collected from all participants at the defined time period in the morning (9:30-10:30) to avoid diurnal variation of saliva flow. Sample collections were performed using Salivettes*®* or Salikids*®* (Sarstedt AG & Co., KG, Numbrecht, Germany). The kit comprises a plastic coat tube and a sponge roll attached with a string for preventing aspiration. The sponge roll was placed under the tongue for 2 minutes to impregnate with a sufficient amount of saliva. Immediately after removing from the mouth, the sponge roll was inserted in a coat tube to cool on ice. The tube was centrifuged at 3,000 x g for 5 minutes at 5 °C to yield clear supernatant fluid (testing saliva sample). The testing saliva samples thus obtained were dispensed into several portions. One hundred μl of the sample was immediately mixed with 500 μl of inactivation solution for a SARS-CoV-2 PCR test to confirm test-negative. The residual samples were stored at −80 °C. Blood samples were collected from an antecubital vein and allowed to stand for 15 minutes at room temperature. The blood tubes were centrifuged at 2,000 x g for 15 minutes. The resultant serum samples were stored at −20 °C until they were transported to the assay laboratory in Kanagawa Dental University where all samples were collected, stored at −80 °C and immunologically analyzed.

### Detection of anti-SARS-CoV-2 antibodies

Enzyme-linked immunosorbent assays (ELISAs) to measure the binding of IgA in saliva, as well as IgA and IgG in serum, to SARS-CoV-2 spike-1 were performed using an assay system modifying the human IgA ELISA quantitation set (#88-102; Bethyl Laboratories, Montgomery, Texas, USA) that had been reported by Yamamoto et al (61). In this kit, anti-IgA or anti-IgG antibodies are adsorbed on the plate as capture antibodies beforehand. For the measurement of IgA in saliva, spike-1-mFc recombinant protein (#40591-V05H1; Sino Biological, Beijin, China) was used as the SARS-CoV-2 antigen based on previous reports (39). However, since that antigen reacts with IgG, spike-1-His recombinant protein (#40591-V08B1; Sino Biological, Beijin, China) was used to measure the cross-antibodies in serum. Both antigens are SARS-CoV-2 spike 1 subunit proteins containing RBD. Biotin-labeling of the antigens was performed using a labeling kit (#BK01; Dojindo Laboratories, Kumamoto, Japan) according to the manufacturer’s instructions. Half-well plates added with saliva samples at 1:500 dilution or serum samples at 1:100 dilution in carbonate-bicarbonate buffer were incubated for 1 hour at 25 °C, and then washed with washing buffer. To the wells thus coated with diluted saliva or serum samples for measurement of SARS-CoV-2-reactive IgA or IgG antibodies, 100 μl per well of biotin-labeled antigen at a concentration of 1 μg/ml was added, and incubated for 1 hour at 25 °C. Wells were washed five times with washing buffer, and then plates were incubated with horseradish peroxidase (HRP) solution A (Bethyl laboratories) for 1hour at 25 °C. Plates were developed by addition of the HRP substrate, TMB (Bethyl Laboratories, Inc., Waltham, Massachusetts, USA), for 15 minutes at 25 °C. Then the developing reaction was quenched by adding stop solution.

ODs were measured at 450 nm in a microplate absorbance reader (Bio-Rad Laboratories, Hercules, CA, USA). As positive control, commercially available two antibody products, i.e. spike-neutralizing IgG antibody (cat#40592-R001, Sino Biological; from 0 to 20 μg/ml) and spike-neutralizing IgA antibody (cat#E-AB-V1027, Elabscience, Houston, Texas, USA; from 0 to 2 μg/ml) for detecting SARS-CoV-2-reacting IgG and IgA, respectively. A positive control and a negative control PBS were added to every assay plate for validation. Background absorbance value for negative control was subtracted from absorbance value for each saliva or serum sample to account for non-specific binding of biotin-labeled antigen to wells without antibody. The detection limits for saliva IgA, serum IgA and serum IgG were 0.002, 0.163 and 1.0 ng/ml, respectively; thus, the tests for saliva IgA, serum IgA and serum IgG were considered positive when the values observed were equal to or above the detection limits.

### Questionnaire

At saliva collection with or without blood drawing, participants were asked to complete a questionnaire containing: current demographic characteristics (e.g. age, gender, body-mass index [BMI], nativity, physical health, comorbidities such as pollinosis and other allergic disorders), oral care (particularly, daily frequency of tooth-brushing), history of vaccination for any viral diseases (e.g. influenza, type-B viral hepatitis, rubella and mumps), current medication and dietary habits including consumption of fermented foods and/or supplementary diets, as well as COVID-19-related symptoms.

### Statistics

Comparison of positivity for SARS-CoV-2-reactive Igs among different participant groups was analyzed using the chi-squared test. The correlation between saliva IgA, serum IgA, and serum IgG concentrations reactive to SARS-CoV-2 was evaluated by Pearson’s correlation coefficient. The results with P-values less than 0.05 were considered statistically significant. Analyses were performed using IBM SPSS Statistics version 27 (IBM, USA).

**Supplementary Materials**

## Data Availability

All data associated with this study are present in the paper.

## Acknowledgments

We thank the patients who consented to participate in this study. We are also grateful to Makiko Yamada of the Research Support Center, Graduate School of Dentistry, Kanagawa Dental College, for her invaluable technical assistance.

## Funding

K.T. was supported for this work by the EPS Holdings Inc.

## Author contributions

Conceptualization: K. Tsukinoki, T. Yamamoto

Methodology: K. Tsukinoki

Investigation: W. Sakaguchi,

Sample collection: J. Saito, K. Iguchi, Y. Inoue, S. Ishii, C. Sato, M. Yokoyama, Y. Shiraishi, N. Kato, A. Makabe, A. Saito

Funding acquisition: T. Yamamoto

Analysis of data: H. Shimada

Project administration: K. Tsukinoki, T. Yamamoto

Supervision: M. Tanji, I. Nagaoka, J. Saruta, T. Yamaguchi, S. Kimoto Writing: H.Yamaguchi

## Competing interests

As employees of EPS, the following people are paid by EPS; T. Yamamoto, Y. Shiraishi, N. Kato, H. Shimada, A. Makabe, A. Saito, M. Tanji. Other authors declare no conflict of interest.

## Data and materials availability

All data associated with this study are present in the paper.

## Notes

### Clinical Protocols

https://center6.umin.ac.jp/cgi-open-bin/ctr/ctr_view.cgi?recptno=R000049910

### Funding Statement

Keiichi Tsukinoki was supported for this work by the EPS Holdings Inc.

### Author Declarations

Approval to undertake the study was obtained from the Kanagawa Dental University Research Ethics Review Board (approval number: 792) on April 6, 2021. The study adhered to all the guidelines set forth in the Declaration of Helsinki (amended in October 2013), the Ethical Guidelines for Medical Research in Humans (noticed by the Ministry of Health, Labor and Welfare, the Japanese Government, in December 2014 [amended in February 2017]) and the Guidance of the Ethical Guidelines for Medical Research in Humans (enacted by the Ministry of Education and Science and the Ministry of Health, Labor and Welfare, the Japanese Government, in February 2015 [amended in March 2017]). Written informed consent was obtained from all participants themselves or informed assent from their legal representatives, mostly parents (in the case of children or adolescent participants aged below 20 years), prior to the study onset. The study was registered on April 1, 2021 in the database of the University Hospital Medical Information Network Clinical Trial Registry (UMIN-CTR) that meets the JCMJE standards. The study ID was UMIN000043717.

### Summary of Updates

The author's e-mail address was misspelled and has been corrected.

## References and Notes

1. N. Zhu, D. Zhang, W. Wang, X. Li, B. Yang, J. Song, X. Zhao, B. Huang, W. Shi, R. Lu, P. Niu, F. Zhan, X. Ma, D. Wang, W. Xu, G. Wu, G. F. Gao, D.Phil., W. Tan, for the China Novel Coronavirus Investigating and Research Team, A novel coronavirus from patients with pneumonia in China. 2019. N. Engl. J. Med. 382, 727–733 (2020)

2. World Health Organization, Coronavirus disease (COVID-19) weekly epidemiological update - 3 November 2020. [Internet] https://www.who.int/publications/m/item/weekly-epidemiological-update-november-20203

3. K. O. Kwok, F. Lai, W. I. Wei, S. Y. S. Wong, J. W. T. Tang, Herd immunity - estimating the level required to halt the COVID-19 epidemics in affected countries. J. Infect. 80, e32–33 (2020)

4. Johns Hopkins University and medicine, coronavirus resource center, COVID-19 Dashboard by the Center for Systems Science and Engineering (CSSE) at Johns Hopkins University (JHU). [Internet] https://www.arcgis.com/apps/dashboards/bda7594740fd40299423467b48e9ecf6

5. S. E. Butler, A. R. Crowley, H. Natarajan, S. Xu, J. A. Weiner, C. A. Bobak, D. E. Mattox, J. Lee, W. Wieland-Alter, R. I. Connor, P. F. Wright, M. E. Ackerman, Distinct features and functions of systemic and mucosal humoral immunity among SARS-CoV-2 convalescent individuals. Front. Immunol. 11, 618685 (2021)

6. D. Sterlin, A. Mathian, M. Miyara, A. Mohr, F. Anna, L. Claër, P. Quentric, J. Fadlallah, H. Devilliers, P. Ghillani, C. Gunn, R. Hockett, S. Mudumba, A. Guihot, C. Luyt, J. Mayaux, A. Beurton, S. Fourati, T. Bruel, O. Schwartz, J. Lacorte, H. Yssel, C. Parizot, K. Dorgham, P. Charneau, Z. Amoura, G. Gorochov, IgA dominates the early neutralizing antibody response to SARS-CoV-2. Sci. Transl. Med. 13, eabd2223 (2021)

7. A. Varadhachary, D. Chatterjee, J. Garza, R. P. Garr, C. Foley, A. Letkeman, J. Dean, D. Haug, J. Breeze, R. Traylor, A. Malek, R. Nath, L. Linbeck III, Salivary anti-SARS-CoV-2 IgA as an accessible biomarker of mucosal immunity against COVID-19. medRxiv, doi:10.1101/2020.08.07.20170258 (2020)

8. Z. Wang, J. C. C. Lorenzi, F. Muecksch, S. Finkin, C. Viant, C. Gaebler, M. Cipolla, H. Hoffman, T. Y. Oliveira, D. A. Oren, V. Ramos, L. Nogueira, E. Michailidis, D. F. Robbiani Gazumyan, C. M. Rice, T. Hatziioannou, P. D. Bieniasz, M. Caskey, M. C. Nussenzweig, Enhanced SARS-CoV-2 neutralization by secretory IgA in vitro. bioRxiv, doi: 10.1101/2020.09.09.288555 (2020)

9. K. W. Ng, N. Faulkner, G. H. Cornish, A. Rosa, R. Harvey, S. Hussain, R. Ulferts, C. Earl, A. G. Wrobel, D. J. Benton, C. Roustan, W. Bolland, R. Thompson, A. Agua-Doce, P. Hobson, J. Heaney, H. Rickman, S. Paraskevopoulou, C. F. Houlihan, K. Thomson, E. Sanchez, G. Y. Shin, M. J. Spyer, D. Joshi, N. O’Reilly, P. A. Walker, S. Kjaer, A. Riddell, C. Moore, B. R. Jebson, M. Wilkinson, L. R. Marshall, E. C. Rosser, A. Radziszewska, H. Peckham, C. Ciurtin, L. R. Wedderburn, R. Beale, C. Swanton, S. Gandhi, B. Stockinger, J. McCauley, S. J. Gamblin, L. E. McCoy, P. Cherepanov, E. Nastouli, G. Kassiotis, Preexisting and de novo humoral immunity to SARS-CoV-2 in humans. Science 370, 1339–1343 (2020)

10. P. P. Jonard, J. C. Rambaud, C. Dive, J. P. Vaerman, A. Galian, D. L. Delacroix, Secretion of immunoglobulins and plasma proteins from the jejunal mucosa. Transport rate and origin of polymeric immunoglobulin A. J. Clin. Invest. 74, 525–535 (1984)

11. P. Brandtzaeg, Mucosal immunity: induction, dissemination, and effector functions. Scand. J. Immunol. 70, 505–515 (2009)

12. S. Krugmann, R. J. Pleass, J. D. Atkin, J. M. Woof, Structural requirements for assembly of dimeric IgA probed by site-directed mutagenesis of J chain and a cysteine residue of the alpha-chain CH2 domain. J. Immunol. 159, 244–249 (1997)

13. C. S. Kaetzel, The polymeric immunoglobulin receptor: bridging innate and adaptive immune responses at mucosal surfaces. Immunol. Rev. 206, 83–99 (2005).

14. J.M. Woof, M.W. Russell, Structure and function relationships in IgA. Mucosal Immunol. 4, 590–597 (2011)

15. M. A. Kerr, The structure and function of human IgA. Biochem. J. 271, 285–296 (1990)

16. J. F. Bermejo-Martin, M. González-Rivera, R. Almansa, D. Micheloud, A. P. Tedim, M. Domínguez-Gil, S. Resino, M. Martín-Fernández, P. R. Murua, F. Pérez-García, L. Tamayo, R. Lopez-Izquierdo, E. Bustamante, C. Aldecoa, J. M. Gómez, J. Rico-Feijoo, A. Orduña, R. Méndez, I. F. Natal, G. Megías, M. González-Estecha, D. Carriedo, C. Doncel, N. Jorge, A. Ortega, A. de la Fuente, F. del Campo, J.A. Fernández-Ratero, W. Trapiello, P. González-Jiménez, G. Ruiz, A. A. Kelvin, A. T. Ostadgavahi, R. Oneizat, L. M. Ruiz, I. Miguéns, E. Gargallo, I. Muñoz, S. Pelegrin, S. Martín, P. G. Olivares, J. A. Cedeño, T. R. Albi, C. Puertas, J.Á. Berezo, G. Renedo, R. Herrán, J. Bustamante-Munguira, P. Enríquez, R. Cicuendez, J. Blanco, J. Abadia, J. G. Barquero, N. Mamolar, N. Blanca-López, L. J. Valdivia, B. F. Caso, M.Á. Mantecón, A. Motos, L. Fernandez-Barat, R. Ferrer, F. Barbé, A. Torres, R. Menéndez, J. M. Eiros, D. J. Kelvin, Viral RNA load in plasma is associated with critical illness and a dysregulated host response in COVID-19. Crit. Care 24, 691 (2020)

17. M. W. Russell, Z. Moldoveanu, P. L. Ogra, J. Mestecky, Mucosal immunity in COVID-19: a neglected but critical aspect of SARS-CoV-2 infection. Front. Immunol. 11, 611337 (2020)

18. N. Huang, P. Pérez, T. Kato, Y. Mikami, K. Okuda, R. C. Gilmore, C. D. Conde, B. Gasmi, S. Stein, M. Beach, E. Pelayo, J. O. Maldonado, B. A. Lafont, S. Jang, N. Nasir, R. J. Padilla, V. Murrah, R. Maile, W. Lovell, S. M. Wallet, N. M. Bowman, S. L. Meinig, M. C. Wolfgang, S. N. Choudhury, M. Novotny, B. D. Aevermann, R. H. Scheuermann, G. Cannon, C. W. Anderson, R. E. Lee, J. T. Marchesan, M. Bush, M. Freire, A. J. Kimple, D. L. Herr, J. Rabin, A. Grazioli, S. Das, B. N. French, T. Pranzatelli, J. A. Chiorini, D. E. Kleiner, S. Pittaluga, S. M. Hewitt, P. D. Burbelo, D. Chertow, NIH COVID-19 Autopsy Consortium, HCA Oral and Craniofacial Biological Network, K. Frank, J. Lee, R. C. Boucher, S. A. Teichmann, B. M. Warner, K. M. Byrd, SARS-CoV-2 infection of the oral cavity and saliva. Nat. Med. 27, 892-903 (2021)

19. M. Hoffmann, H. Kleine-Weber, S. Schroeder, N. Krüger, T. Herrler, S. Erichsen, T. S. Schiergens, G. Herrler, N. Wu, A. Nitsche, M. A. Müller, C. Drosten, S. Pöhlmann, SARSCoV-2 cell entry depends on ACE2 and TMPRSS2 and is blocked by a clinically proven protease inhibitor. Cell 181, 271–280 (2020)

20. L. Chen, J. Zhao, J. Peng, X. Li, X. Deng, Z. Geng, Z. Shen, F. Guo, Q. Zhang, Y. Jin, L. Wang, S. Wang, Detection of SARS-CoV-2 in saliva and characterization of oral symptoms in COVID-19 patients. Cell Prolif. 53, e12923 (2020)

21. N. L. Bert, A. T. Tan, K. Kunasegaran, C. Y. L. Tham, M. Hafezi, A. Chia, M. H. Y. Chng, M. Lin, N. Tan, M. Linster, W. N. Chia, M. I. Chen, L. Wang, E. E. Ooi, S. Kalimuddin, P. A. Tambyah, J. G. Low, Y. Tan, A. Bertoletti, SARS-CoV-2-specific T cell immunity in cases of COVID-19 and SARS, and uninfected controls. Nature 584, 457–462 (2020)

22. J. Braun, L. Loyal, M. Frentsch, D. Wendisch, P. Georg, F. Kurth, S. Hippenstiel, M. Dingeldey, B. Kruse, F. Fauchere, E. Baysal, M. Mangold, L. Henze, R. Lauster, M. A. Mall, K. Beyer, J. Röhmel, S. Voigt, J. Schmitz, S. Miltenyi, I. Demuth, M. A. Müller, A. Hocke, M. Witzenrath, N. Suttorp, F. Kern, U. Reimer, H. Wenschuh, C. Drosten, V. M. Corman, C. Giesecke-Thiel, L. E. Sander, A. Thiel, SARS-CoV-2-reactive T cells in healthy donors and patients with COVID-19. Nature 587, 270–274 (2020)

23. M. S. Maddur, S. Lacroix-Desmazes, J. D. Dimitrov, M. D. Kazatchkine, J. Bayry, S. V. Kaveri, Natural antibodies: from first-line defense against pathogens to perpetual immune homeostasis. Clin. Rev. Allergy Immunol. 58, 213–228 (2020)

24. N. Sood, P. Simon, P. Ebner, D. Eichner, J. Reynolds, E. Bendavid, J. Bhattacharya, Seroprevalence of SARS-CoV-2-specific antibodies among adults in Los Angeles County, California, on April 10-11, 2020. JAMA 323, 2425–2427 (2020)

25. M. C. Dalakas, K. Bitzogli, H. Alexopoulos, Anti-SARS-CoV-2 antibodies within IVIg preparations: cross-reactivities with seasonal coronaviruses, natural autoimmunity, and therapeutic implications. Front. Immunol. 12, 627285 (2021)

26. M. Ejemel, Q. Li, S. Hou, Z. A. Schiller, J. A. Tree, A. Wallace, A. Amcheslavsky, N. K. Yilmaz, K. R. Buttigieg, M. J. Elmore, K. Godwin, N. Coombes, J. R. Toomey, R. Schneider, S. Ramchetty, B. J. Close, D. Chen, H. L. Conway, M. Saeed, C. Ganesa, M. W. Carroll, L. A. Cavacini, M. S. Klempner, C. A. Schiffer, Y. Wang, A cross-reactive human IgA monoclonal antibody blocks SARS-CoV-2 spike-ACE2 interaction. Nat. Commun. 11, 4198 (2020).

27. S. E. Faustini, S. E. Jossi, M. Perez-Toledo, A. M. Shields, J. D. Allen, Y. Watanabe, M. L. Newby, A. Cook, C. R. Willcox, M. Salim, M. Goodall, J. L. Heaney, E. Marcial-Juarez, G. L. Morley, B. Torlinska, D. C. Wraith, T. V. Veenith, S. Harding, S. Jolles, M. J. Ponsford, T. Plant, A. Huissoon, M. K. O’Shea, B. E. Willcox, M. T. Drayson, M. Crispin, A. F. Cunningham, A. G. Richter, Detection of antibodies to the SARS-CoV-2 spike glycoprotein in both serum and saliva enhances detection of infection. medRxiv, doi: 10.1101/2020.06.16.20133025 (2020)

28. N. Pisanic, P. R. Randad, K. Kruczynski, Y. C. Manabe, D. L. Thomas, A. Pekosz, S. L. Klein, M. J. Betenbaugh, W. A. Clarke, O. Laeyendecker, P. P. Caturegli, H. B. Larman, B. Detrick, J. K. Fairley, A. C. Sherman, N. Rouphael, S. Edupuganti, D. A. Granger, S. W. Granger, M. H. Collins, C. D. Heaney, COVID-19 serology at population scale: SARS-CoV-2-specific antibody responses in saliva. J. Clin. Microbiol. 59, e02204–02220 (2020)

29. Q. Jing, M. Liu, Z. Zhang, L. Fang, J. Yuan, A. Zhang, N. E. Dean, L. Luo, M. Ma, I. Longini, E. Kenah, Y. Lu, Y. Ma, N. Jalali, Z. Yang, Y. Yang, Household secondary attack rate of COVID-19 and associated determinants in Guangzhou, China: a retrospective cohort study. Lancet Infect. Dis. 20, 1141–1150 (2020)

30. J. Zhang, M. Litvinova, Y. Liang, Y. Wang, W. Wang, S. Zhao, Q. Wu, S. Merler, C. Viboud Vespignani, M. Ajelli, H. Yu, Changes in contact patterns shape the dynamics of the COVID-19 outbreak in China. Science 368, 1481–1486 (2020)

31. W. Li, B. Zhang, J. Lu, S. Liu, Z. Chang, C. Peng, X. Liu, P. Zhang, Y. Ling, K. Tao, J. Chen, Characteristics of household transmission of COVID-19. Clin. Infect. Dis. 71, 1943–1946 (2020)

32. E. S. Rosenberg, E. M. Dufort, D. S. Blog, E. W. Hall, D. Hoefer, B. P. Backenson, A. T. Muse, J. N. Kirkwood, K. S. George, D. R. Holtgrave, B. J. Hutton, H. A. Zucker, for the New York State Coronavirus 2019 Response Team, COVID-19 testing, epidemic features, hospital outcomes, and household prevalence, New York State—March 2020. Clin. Infect. Dis. 71, 1953–1959 (2020)

33. J. Wu, Y. Huang, C. Tu, C. Bi, Z. Chen, L. Luo, M. Huang, M. Chen, C. Tan, Z. Wang, K. Wang, Y. Liang, J. Huang, X. Zheng, J. Liu, Household transmission of SARS-CoV-2, Zhuhai, China, 2020. Clin. Infect. Dis. 71, 2099–2108 (2020)

34. A. R. Yousaf, L. M. Duca, V. Chu, H. E. Reses, M. Fajans, E. M. Rabold, R. L. Laws, R. Gharpure, A. Matanock, A. Wadhwa, M. Pomeroy, H. Njuguna, G. Fox, A. M. Binder, A. Christiansen, B. Freeman, C. Gregory, C. H. Tran, D. Owusu, D. Ye, E. Dietrich, E. Pevzner, E. E. Conners, I. Pray, J. Rispens, J. Vuong, K. Christensen, M. Banks, M. O’Hegarty, L. Mills, S. Lester, N. J. Thornburg, N. Lewis, P. Dawson, P. Marcenac, P. Salvatore, R. J. Chancey, V. Fields, S. Buono, S. Yin, S. Gerber, T. Kiphibane, T. Dasu, S. Bhattacharyya, R. Westergaard, A. Dunn, A. J. Hall, A. M. Fry, J. E. Tate, H. L. Kirking, S. Nabity, A prospective cohort study in nonhospitalized household contacts with severe acute respiratory syndrome coronavirus 2 infection: symptom profiles and symptom change over time. Clin. Infect. Dis. 73, e1841–1849 (2021)

35. Q. Bi, Y. Wu, S. Mei, C. Ye, X. Zou, Z. Zhang, X. Liu, L. Wei, S. A Truelove, T. Zhang, W. Gao, C. Cheng, X. Tang, X. Wu, Y. Wu, B. Sun, S. Huang, Y. Sun, J. Zhang, T. Ma, J. Lessler, T. Feng, Epidemiology and transmission of COVID-19 in 391 cases and 1286 of their close contacts in Shenzhen, China: a retrospective cohort study. Lancet Infect. Dis. 20, 911–919 (2020)

36. L. Luo, D. Liu, X. Liao, X. Wu, Q. Jing, J. Zheng, F. Liu, S. Yang, H. Bi, Z. Li, J. Liu, W. Song, W. Zhu, Z. Wang, X. Zhang, Q. Huang, P. Chen, H. Liu, X. Cheng, M. Cai, P. Yang, X. Yang, Z. Han, J. Tang, Y. Ma, C. Mao, Contact settings and risk for transmission in 3410 close contacts of patients with COVID-19 in Guangzhou, China: a prospective cohort study. Ann. Intern. Med. 173, 879–887 (2020)

37. H. C. Maltezou, R. Vorou, K. Papadima, A. Kossyvakis, N. Spanakis, G. Gioula, M. Exindari, S. Metallidis, A. N. Lourida, V. Raftopoulos, E. Froukala, B. Martinez-Gonzalez, A. Mitsianis, E. Roilides, A. Mentis, A. Tsakris, A. Papa, Transmission dynamics of SARS-CoV-2 within families with children in Greece: a study of 23 clusters. J.Med.Virol. 93, 1414–1420 (2021)

38. S. Kurup, R. Burgess, F. Tine, A. Chahroudi, D. L. Lee, SARS-CoV-2 infection and racial disparities in children: protective mechanisms and severe complications related to MIS-C. J. Racial Ethn. Health Disparities, 13, 1–7 (2021)

39. K. Tsukinoki, T. Yamamoto, K. Handa, M. Iwamiya, J. Saruta, S. Ino, T. Sakurai, Detection of cross-reactive immunoglobulin A against the severe acute respiratory syndrome-coronavirus-2 spike 1 subunit in saliva. PLoS ONE 16, e0249979 (2021).

40. A. Jafarzadeh, M. Sadeghi, G. A. Karam, R. Vazirinejad, Salivary IgA and IgE levels in healthy subjects: relation to age and gender. Braz. Oral Res. 24, 21–27 (2010)

41. I. D. Miletic, S. S. Schiffman, V. D. Miletic, E. A. Sattely-Miller, Salivary IgA secretion rate in young and elderly persons. Physiol. Behav. 60, 243–248 (1996)

42. J. Kugler, M. Hess, D. Haake, Secretion of salivary immunoglobulin A in relation to age, saliva flow, mood states, secretion of albumin, cortisol, and catecholamines in saliva. J. Clin. Immunol. 12, 45–49 (1992)

43. A. Banzhoff, A. Dulleck, S. Petzoldt, C. H. Rieger, Salivary anti-RSV IgA antibodies and respiratory infections during the first year of life in atopic and non-atopic infants. Pediatr. Allergy Immunol. 5, 46–52 (1994)

44. B. Hewson-Bower, P. D. Drummond, Secretory immunoglobulin A increases during relaxation in children with and without recurrent upper respiratory tract infections. J. Dev. Behav. Pediatr. 17, 311–316 (1996)

45. P. D. Drummond, B. Hewson-Bower, Increased psychosocial stress and decreased mucosal immunity in children with recurrent upper respiratory tract infections. J. Psychosom. Res. 43, 271–278 (1997)

46. Y. Yodfat, I. Silvian, A prospective study of acute respiratory tract infections among children in a kibbutz: the role of secretory IgA and serum immunoglobulins. J. Infect. Dis. 136, 26–30 (1977).

47. C. M. Stover, Mechanisms of stress-mediated modulation of upper and lower respiratory tract infections. Adv. Exp. Med. Biol. 874, 215–223 (2016)

48. M. Hess, J. Kugler, D. Haake, J. Lamprecht, Reduced concentration of secretory IgA indicates changes of local immunity in children with adenoid hyperplasia and secretory otitis media. ORL J. Otorhinolaryngol. Relat. Spec. 53, 339–341 (1991)

49. S. Chaushu, E. Yefenof, A. Becker, J. Shapira, G. Chaushu, A link between parotid salivary Ig level and recurrent respiratory infections in young Down’s syndrome patients. Oral Microbiol. Immunol. 17, 172–176 (2002)

50. M. Gleeson, W. A. McDonald, D. B. Pyne, A. W. Cripps, J. L. Francis, P. A. Fricker, R. L. Clancy, Salivary IgA levels and infection risk in elite swimmers. Med. Sci. Sports Exerc. 31, 67–73 (1999)

51. A. M. P. Novas, D. G. Rowbottom, D. G. Jenkins, Tennis, incidence of URTI and salivary IgA. Int. J. Sports Med. 24, 223–229 (2003)

52. V. Neville, M. Gleeson, J. P. Folland, Salivary IgA as a risk factor for upper respiratory infections in elite professional athletes. Med. Sci. Sports Exerc. 40, 1228–1236 (2008)

53. M. Gleeson, N. Bishop, M. Oliveira, T. McCauley, P. Tauler, A. S. Muhamad, Respiratory infection risk in athletes: association with antigen-stimulated IL-10 production and salivary IgA secretion. Scand. J. Med. Sci. Sports, 22, 410–417 (2012)

54. E. Tiollier, D. Gomez-Merino, P. Burnat, J. C. Jouanin, C. Bourrilhon, E. Filaire, C. Y. Guezennec, M. Chennaoui, Intense training: mucosal immunity and incidence of respiratory infections. Eur. J. Appl. Physiol. 93, 421–428 (2005)

55. D. Nakamura, T. Akimoto, S. Suzuki, I. Kono, Daily changes of salivary secretory immunoglobulin A and appearance of upper respiratory symptoms during physical training. J. Sports Med. Phys. Fitness, 46, 152–157 (2006)

56. S. G. Byars, S. C. Stearns, J. J. Boomsma, Association of long-term risk of respiratory, allergic, and infectious diseases with removal of adenoids and tonsils in childhood. JAMA Otolaryngol. Head Neck Surg. 144, 594–603 (2018)

57. P. Brandtzaeg, Molecular and cellular aspects of the secretory immunoglobulin system. APMIS, 103, 1–19 (1995)

58. P. Brandtzaeg, F. Johansen, Mucosal B cells: phenotypic characteristics, transcriptional regulation, and homing properties. Immunol. Rev. 206, 32–63 (2005)

59. C. P. Quan, A. Berneman, R. Pires, S. Avrameas, J. P. Bouvet, Natural polyreactive secretory immunoglobulin A autoantibodies as a possible barrier to infection in humans. Infect. Immun. 65, 3997–4004 (1997)

60. Our World in Data, Coronavirus (COVID-19) Vaccinations. [Internet] https://ourworldindata.org/covid-vaccinations

61. Y. Yamamoto, J. Saruta, T. Takahashi, M. To, T. Shimizu, T. Hayashi, T. Morozumi, N. Kubota, Y. Kamata, S. Makino, H. Kano, J. Hemmi, Y. Asami, T. Nagai, K. Misawa, S. Kato, K. Tsukinoki, Effect of ingesting yogurt fermented with Lactobacillus delbrueckii ssp. bulgaricus OLL1073R-1 on influenza virus-bound salivary IgA in elderly residents of nursing homes: a randomized controlled trial. Acta Odontologica Sacnd. 77, 517–524 (2019)

